# Research on CNN-based Models Optimized by Genetic Algorithm and Application in the Diagnosis of Pneumonia and COVID-19

**DOI:** 10.1101/2020.04.21.20072637

**Authors:** Zihan Zeng, Bo Wang, Zhiwen Zhao

## Abstract

In this research, an optimized deep learning method was proposed to explore the possibility and practicality of neural network applications in medical imaging. The method was used to achieve the goal of judging common pneumonia and even COVID-19 more effectively. Where, the genetic algorithm was taken advantage to optimize the Dropout module, which is essential in neural networks so as to improve the performance of typical neural network models. The experiment results demonstrate that the proposed method shows excellent performance and strong practicability in judging pneumonia, and the application of advanced artificial intelligence technology in the field of medical imaging has broad prospects.

## INTRODUCTION

After the Deep Learning leader Geoffrey Hinton ended the dark age of artificial intelligence in 2012, artificial intelligence technology, especially neural network technology, has begun to explode. More and more industries have started to use artificial intelligence technology to achieve the purpose of reducing workload and improving efficiency. As a new generation of artificial intelligence technology after machine learning, neural networks have pushed machine cognition to new heights in terms of speech recognition, text processing, image recognition and so on.

With the improvement of people’s material living standards, the incidence of various diseases is also increasing. According to the survey data of the Chinese CDC, compared with the data from 2003 to 2007, the incidence of cancer in China has increased by about 7% (6.8% for men and 7.5% for women) (Chinese Center for Disease Control and Prevention, 2017). According to the World Health Organization’s survey of patients with colon cancer in the United States, the incidence and mortality of colon cancer in the United States have been declining for the past two decades, thanks to the colonoscopy screening vigorously promoted by the United States government. In the zero, one, and two stages of cancer, the cure rate and survival rate of patients are very high, and even in the third stage, the cure rate is as high as 72% (World Health Organization, 2019). The diagnosis of a tumor relies heavily on medical imaging, including X-ray and CT imaging. Similarly, the determination of pneumonia can also help confirm the diagnosis through therapeutic impact technology. Although the diagnosis of pneumonia also includes blood tests and sputum cultures, it is exceptionally efficient to perform medical imaging tests.

According to the “State Council’s Report on the Management of Physician Teams and the Implementation of Practising Physicians,” as of the end of 2018, there were 2.59 doctors per 1,000 population in China (the number of doctors per thousand population in developed countries such as Germany and Austria exceeds 4). The number of public health physicians is inadequate and is decreasing year by year (China National People’s Congress, 2019). Therefore, how to find and diagnose the disease earlier and treat it with the limited number of doctors has become an important research direction of the medical community in China and even in the developing countries.

Artificial intelligence technology takes on this critical mission. At present, the general view in the academic community: the application of artificial intelligence technology in medical imaging can significantly reduce the repetitive work of radiologists, and reduce human errors, thereby improving the diagnosis rate. Since artificial intelligence technology has demonstrated capabilities beyond human cognition in such areas as face recognition and gesture recognition, it is of significant research and practical value whether it is possible to recognize or even detect features that are difficult to find by humans in medical images. For developing countries like China, the practice of artificial intelligence technology in medical imaging can greatly alleviate or even change the medical model of “more patients, fewer doctors” or “two hours in line, five minutes to judge”.

Whether it is facing a persistent disease such as cancer, or a highly contagious and mutating epidemic disease, such as various pneumonias. Artificial intelligence technology can bear corresponding responsibilities and promote the improvement of the medical system. In August 2017, Tencent released “Tencent Miying”, an artificial intelligence medical imaging technology to assist doctors for early screening of esophageal cancer (Tencent, 2018). According to the official Tencent report, before the release, “Tencent Miying” had already cooperated with Shenzhen Nanshan Hospital and Zhongshan Cancer Hospital and had achieved excellent results.

Although the number of papers related to artificial intelligence in China is increasing every year, the number of papers in the field of artificial intelligence medical care is minimal. Most of the papers are concentrated in the fields of natural language processing, real-time image, and video recognition. According to the “2019 China Medical Artificial Intelligence Development Report”, the clinical application fields of artificial medical intelligence in China are mainly concentrated in pediatrics, dermatology, and retinopathy, etc., and there are still some shortcomings in the field of medical imaging (Zhang, 2019).

The United States was one of the first countries to conduct artificial intelligence research. According to data records, the earliest combination of artificial intelligence and medical fields was the University of Leeds, the UK’s top university. The AAPHelp developed by it was the earliest medical artificial intelligence system for the auxiliary diagnosis of severe abdominal pain.

In 2016, Google’s DeepMind participated in research on the treatment of head and neck cancer and cooperated with eye hospitals to apply artificial intelligence technology to the early detection and treatment of eye diseases that threaten vision(Timothy, 2019).

At present, the most well-known in the field of artificial medical intelligence abroad is IBM Watson, which not only involves the screening of tumor images but also can screen historical cancer treatment records and provide reliable treatment solutions. Watson is currently running in the top three areas of cancer treatment (Li, 2019).

In addition to this, Deep Learning scholar Andrew Ng and his team submitted a paper in 2017 and proposed a neural network structure named CheXNet (Rajpurkar, 2017). This neural network structure is used to train a pneumonia image detection classification model, allowing the computer to diagnose pneumonia through chest radiographs automatically. According to the data in the paper, the diagnostic accuracy rate has reached the average level of doctors’ judgment accuracy rate. However, a post by Dr. Luke Oakden-Rayner questioned this result. For the Chest X-ray8 dataset used by Ng’s team, Dr. Luke believes that the labels of this dataset are inaccurate and unclear, which will significantly affect the credibility of the experimental results. The purpose of this topic is to explore the application of neural networks in medical imaging, and to explore the possibility of neural networks replacing doctors to judge diseases. We also provide a part of theory and application basis of “artificial intelligence-medical imaging diagnostic system”. Besides, the current largest open-source COVID-19 CT image dataset was used to explore the feasibility of artificial intelligence technology to diagnose COVID-19.

The innovations of this article are presented below:

- Improved the ordinary Dropout module using genetic algorithms, thereby optimizing the neural network structure;
- The two large data sets are integrated, eliminating the problems caused by inaccurate data label classification of a single data set.

The main work of this paper are as follows:

- Using the optimized InceptionV3 to explore the role of neural networks in judging pneumonia;
- Using the optimized ResNet-152 to explore the role of neural networks in judging pneumonia;
- Using the ResNet-152 to explore the role of neural networks in judging pneumonia caused by the 2019 New Coronavirus;
- Comparing with the latest relevant researches, the experimental results of the proposed algorithms demonstrated in this paper can show good performance.

## DATASET

The dataset used in the main experiment (Judgment of common pneumonia) of this paper integrates two authoritative medical image datasets. The first is the latest Chest X-ray data set (J. et al., 2000), which includes chest X-ray photo collections provided by Guangzhou Maternal and Child Health Center. This dataset was updated again in 2018. The second data set was collected by Shenzhen Third People’s Hospital. Shenzhen Third People’s Hospital is a hospital that focuses on the treatment of infectious diseases and is also a first-line hospital to fight the new coronavirus in 2019 and 2020. The purpose of synthesizing the two data sets is to reduce the adverse effects of the potential errors and omissions of a single data set on the final experiment and to increase the credibility of the final data as much as possible (Join et al., 1938).

Figure 3 shows two examples of chest X-ray images in the dataset. Regular chest X-rays show clear lungs, and there are no abnormally cloudy parts. X-ray imaging of the lungs of patients with pneumonia showed diffuse, cloudy areas.

**Figure 1.**
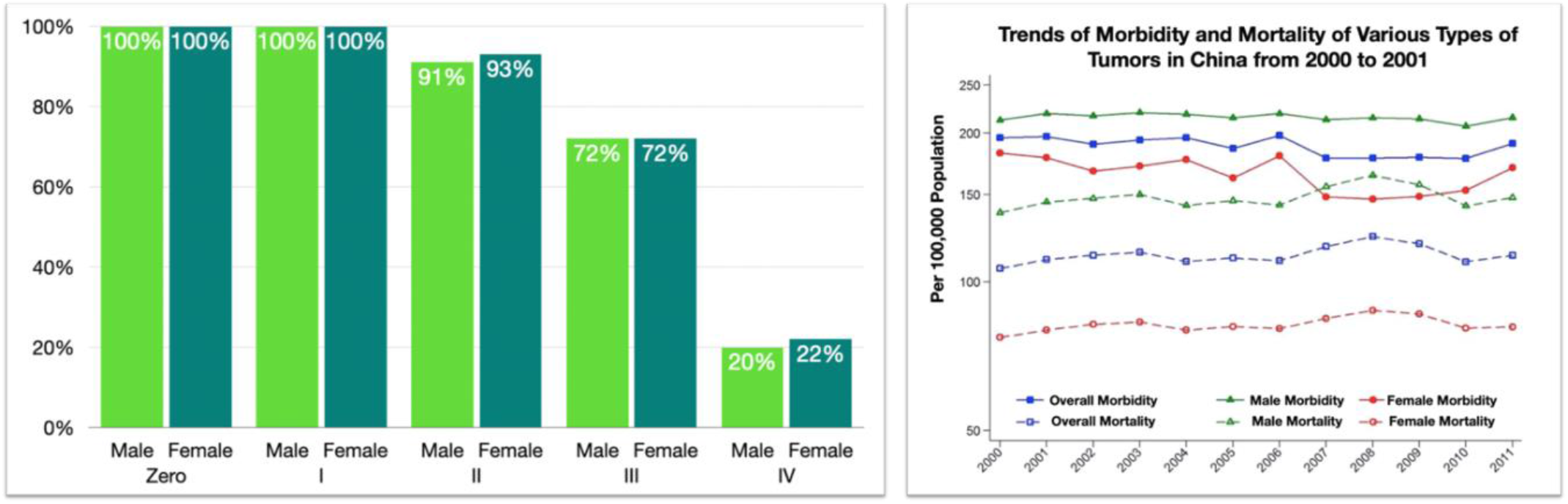
Surveys about Cancer.

**Figure 2.**
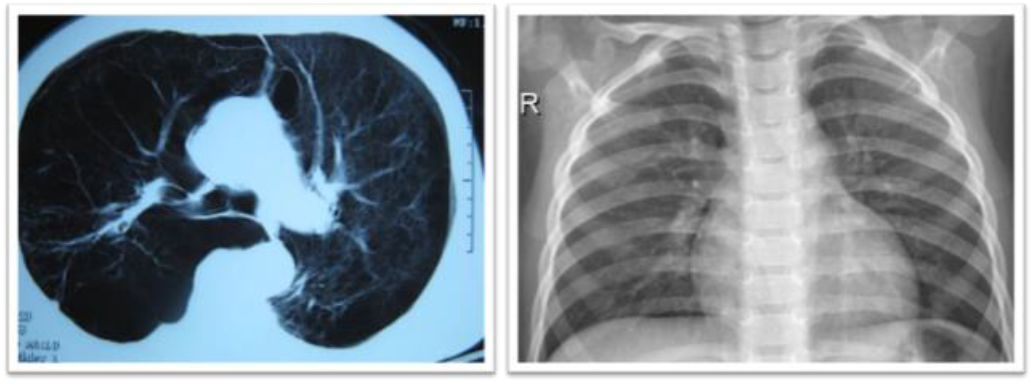
CT Imaging and X-ray Imaging.

**Figure 3.**
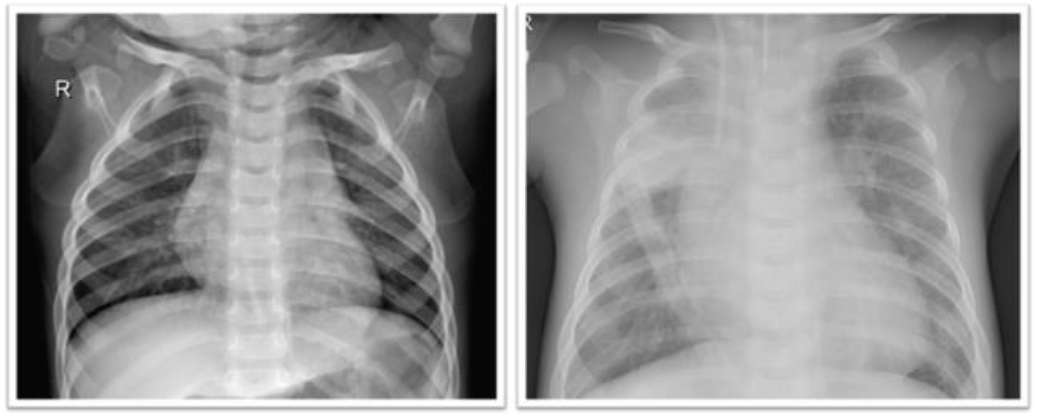
X-rays Image Dataset Examples.

The COVID-19 dataset used was collected by UCSD. In the paper *COVID-CT-Dataset: A CT Scan Dataset about COVID-19*, the researchers extracted CT images from 760 papers which from MedRxiv and BioRxiv related to COVID-19. This dataset is currently the biggest open-source dataset on COVID-19-CT (Zhao, 2020). Random flipping and image cutting methods were used to expand the mounts of the data. Figure 4 are example pictures of the dataset, showing the lung CT images of non-COVID-19 patients and COVID-19 patients, respectively. Since the pictures come from different papers, they need to be processed uniformly during data preprocessing.

**Figure 4.**
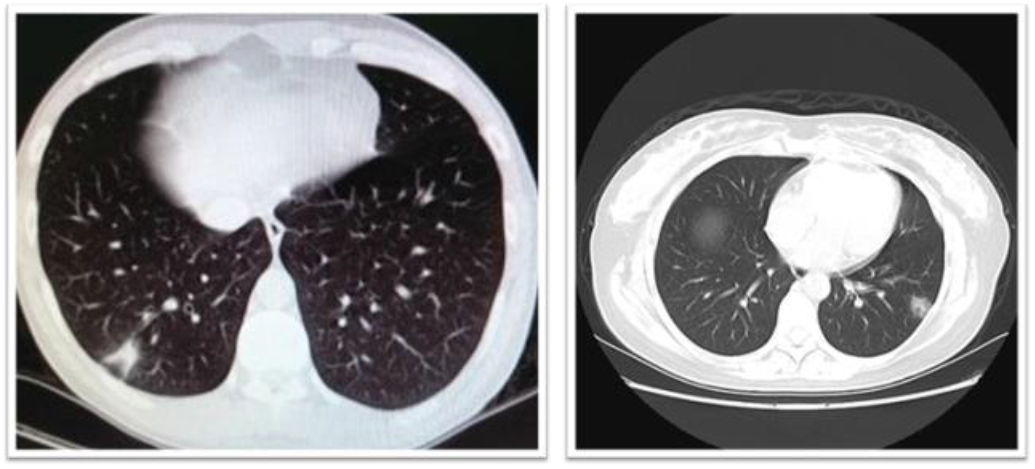
CT Image Dataset Examples.

## METHODS

### Deep Learning Models

Convolutional neural networks have shown significant advantages in image classification (Y et al, 1998; William et al., 2019). With the rapid development of neural networks in the past five years, in order to solve the problem of picture recognition in various situations, convolutional neural networks have developed different branches, and some branches have even iterated four versions. The models used in the experiments in this paper are from the Inception and ResNet neural network families.

InceptionV3, as the third edition of the Inception Deep Learning convolution architecture series, won the first place in the ImageNet Large-scale Visual Recognition Challenge (Jia et al., 2009). Inception V3 not only alleviates the problem of overfitting in the case of limited training data but also conducts a “sparse connection” structure to optimize computing resource allocation and memory footprint. The unique advantage of Inception V3 is that it can extract features in multiple dimensions and retain them. In the field of image recognition, the images in the dataset are very different. Some images have a wider feature distribution, so larger convolution kernels are suitable. For images with more cramped information distribution, smaller convolution kernels can play a better role in feature extraction. However, stacking large convolution kernels will consume many computing resources, and stacking small convolution kernels are prone to overfitting problems, so how to choose a convolution kernel is very important. Inception V3 uses parallel convolution kernels of different sizes to input pictures into these convolution kernels and then performs feature fusion again, which solves the problem of convolution kernel selection (Rohita et al., 2019).

There are many ways of distributing important features in medical images. For example, in lung X-ray imaging, the characterization of lung disease may appear in various parts of the lung, even in several parts. Figure 5 fully confirms this conclusion. Therefore, it can be seen that, for the classification of medical images by neural networks, InceptionV3 can have an excellent effect theoretically.

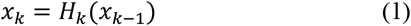

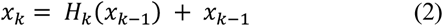

**Figure 5.**
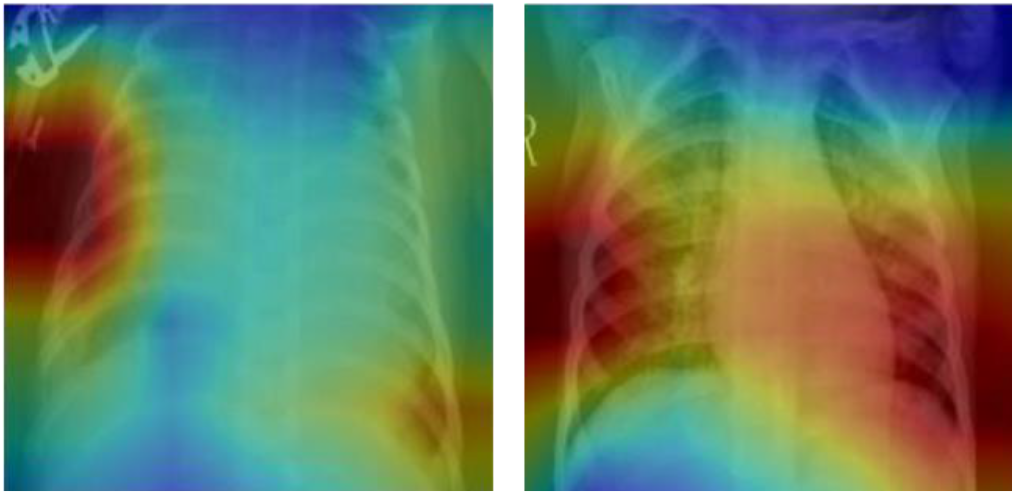
Localization of pneumonia in X-ray Images.

ResNet was published by Dr. Kaiming He in 2016 (Kaiming et al., 2016). Although this neural network structure has a profound number of layers, the problem of deep network degradation and gradient disappearance is well solved by introducing residual units. Equation 1 and Equation 2 are the outputs of the traditional neural network and ResNet at the k-layer, respectively. ResNet’s output at layer *k* adds identity information from the previous layer. The core idea of ResNet is that if the network develops in the direction of degraded performance, it can skip this layer, thereby retaining the best actual performance. ResNet guarantees that while increasing the depth of the network, it can not only converge but also prevent degradation to the greatest extent, thereby ensuring the effectiveness of each network layer. In the field of medical imaging, the research group of Wuhan University used ResNet to study the classification of human protein atlas images (Chang, 2019). The 2016 Deep Learning Methodology and Medical Image Analysis Symposium also exchanged ideas on the use of ResNet for medical image segmentation. Therefore, we believe that the use of ResNet for pneumonia judgment is highly feasible.

### Genetic algorithm optimized Dropout Module

The Dropout function is often used after the convolution layer, and its effect is to randomly discard some features, thereby mitigating the overfitting phenomenon. In general, Dropout has only one parameter *p*, which disables some neurons with probability *p*, so as to achieve the purpose of discarding some features. However, although pure random discarding can alleviate the overfitting phenomenon, there is also a certain probability that it will discard useful features. In my previous research, the Dropout function parameter is generally set to 0.5, which determines whether to discard features like a coin toss (SOR Related et al., 2014).

Admittedly, the Dropout function can well alleviate overfitting and for different neural networks, there could be an optimal parameter value that adapts to the current network structure.

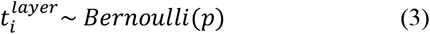

Equation 3 is the most crucial part of the Dropout algorithm. The Bernoulli function uses the parameter *p* to generate a probability vector *t*, each of which is 0 or 1. In a neural network with a Dropout function, after the Dropout function acts on the network of a specific layer, the values of all neuron activation functions in that layer will have a probability of *p* becoming 0, and then stop working. After using the Dropout function, because some neurons lose their effect, the remaining neurons need to be scaled; that is, they need to be multiplied by (1 - *p*).

In the genetic algorithm, individual genes are represented by 0 and 1, and the probability vector in the Dropout algorithm is also 0 or 1. So can consider using the genetic algorithm to optimize the Dropout function and determine the parameter *p* by determining the scaling scale in reverse.

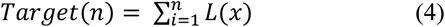

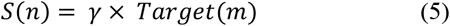

In this part of the experiment, the parameter p is preset to β, and the pre-scale is (1 - *β*). Randomly generate n probability vectors with different scales as the original population {*λ*1, *λ*2, *λ*3,…, *λn*}. The direct goal of the experiment is to find the optimal set of probability vectors, so the objective function can be determined as the sum of the Loss values after randomly extracting some data for training, which can be expressed as Equation 4. In order to better expand the individual differences, a fixed weight *γ* greater than ten can be multiplied based on the objective function to obtain the fitness function *S* (*n*), which is Equation 5. So from a computational point of view, the purpose of the problem is to find the minimum.

Overall, the specific steps of the algorithm are as follows:

1. To preset the parameter *p* as *β*, and preset the breeding cycle *T*, and a small number of training samples are randomly selected;
2. Randomly generate the original population {*λ*1, *λ*2, *λ*3, …, *λn*}
3. To calculate the objective function value *Target* (*n*) and fitness function value *S*(*n*) of the individual;
4. Using the roulette algorithm in the genetic algorithm to select the dominant population and deposit it into the dominant species cluster *G*;
5. Gene exchange and mutation;
6. To determine whether the breeding cycle is over, if it is over, return the optimal solution; if the number of iterations is not completed, continue;
7. After the breeding cycle is over, calculate the proportion of the probability vector with the value of 0 in the optimal solution to get the value *α*;
8. The value of *α* is the optimal value of the parameter *p*.

This method can find an approximate optimal Dropout parameter value, thereby improving the performance of the network as much as possible.

## RESULTS

### Experiment on InceptionV3 with Normal Pneumonia

After the 19th round of training, the decrease rate of the Loss value began to slow down, causing a slight fluctuation, and eventually converging to about 0.02. In order to prevent overfitting, the training automatically stopped. In Figure 6b, the final accuracy of the training set is very high, which is nearly 20% higher than that of the test set. In the subsequent parameters tuning, the overfitting phenomenon still exists. Therefore, it may be because, for the InceptionV3 model, the amount of data in the data pool is not large enough, which makes it impossible to describe the actual distribution of the problem, which leads to overfitting. The overfitting phenomenon in the InceptionV3 experiment also directly caused the AUROC value of this model to be shallow.

**Figure 6.**
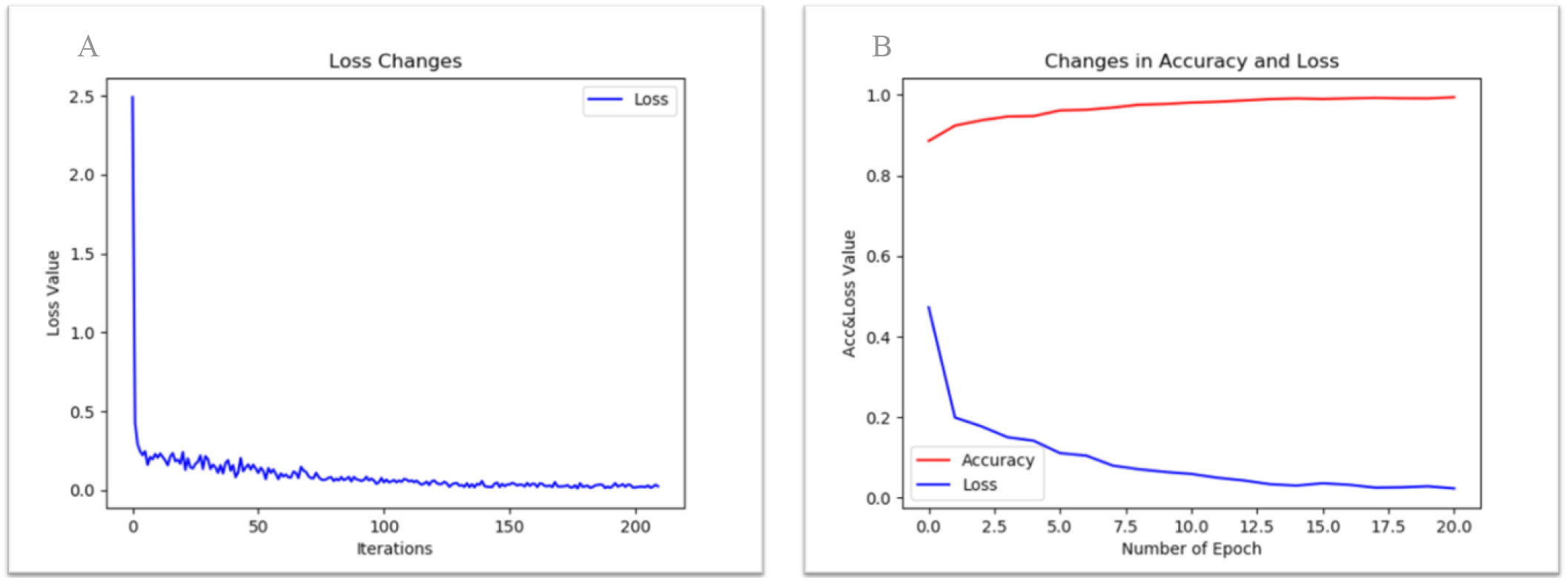
InceptionV3 with Normal Pneumonia Experimental Results. A, Change trend of the loss value about the experiment. It shows the loss value of each iterations. B, Comprehensive change trend of the experiment. It shows the accuracy and the loss value of each epoch.

The accuracy rate after training reached 78.205%. However, the overall results show that the recall rate is high, but the accuracy rate is slightly lower, which means that the model tends to judge most data points as positive examples. From the perspective of F1 scores, the F1 score of InceptionV3 reached 0.85120, which is a good result, proving that the performance of this model has some usability. The AUROC value reflected in the results is only 0.691880341880. According to the data provided in the CheXNet paper (Rajpurkar, 2017), some AUROC value of medical experts in lung disease judgments is greater than 0.7. Therefore, this model does not show much better performance than humans.

### Experiment on ResNet-152 with Normal Pneumonia

Figure 7a shows the complete changes of ResNet-152 experimental Loss. Similar to the InceptionV3 experiment, Loss value also experienced a rapid decline and a stable contraction of the overall network after the first iteration.

**Figure 7.**
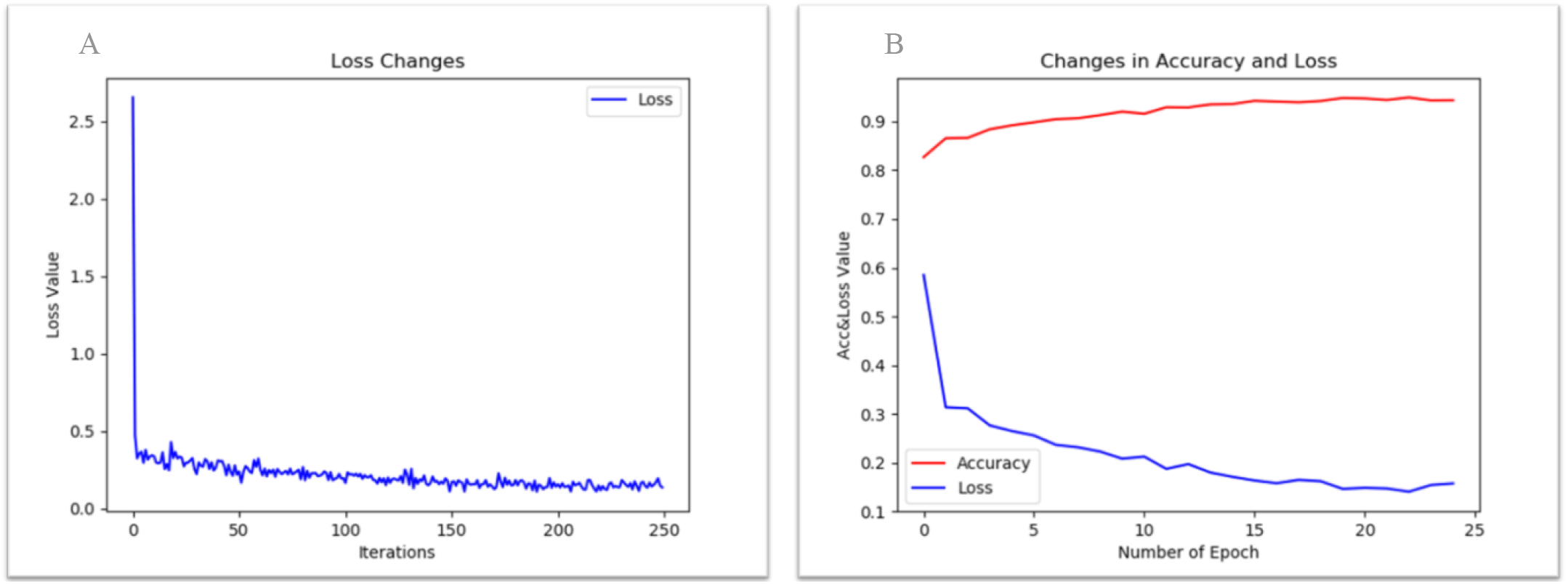
ResNet-152 with Normal Pneumonia Experimental Results. A, Change trend of the loss value about the experiment. It shows the loss value of each iterations. B, Comprehensive change trend of the experiment. It shows the accuracy and the loss value of each epoch.

**Figure 8.**
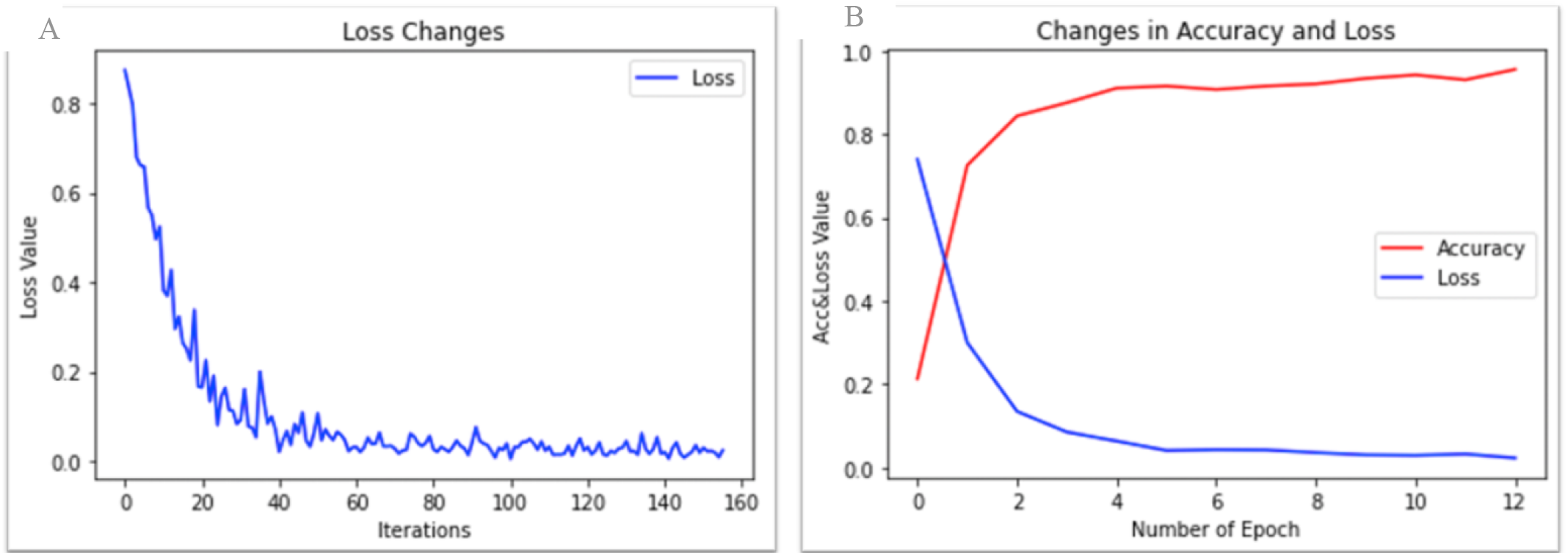
ResNet-152 with COVID-19 Experimental Results. A, Change trend of the loss value about the experiment. It shows the loss value of each iterations. B, Comprehensive change trend of the experiment. It shows the accuracy and the loss value of each epoch.

Due to the increased network depth and increased parameters, the time spent in the ResNet-152 experiment and the number of training rounds are more significant than the InceptionV3 experiment. However, in the ResNet-152 network training, the Loss value did not converge to a minimal value, but it started to produce rebound around 0.14. So in order to prevent overfitting, the training stops automatically.

The experimental results show that the accuracy rate reaches 92.788%, and the recall rate and accuracy rate are both above 0.9, which is an outstanding result. The F1 score also reached 0.94465, which is also a fantastic result, which proves the usability of this model. The AUROC value is 0.908974358974, which also shows that this model is highly likely to have high practicality in medicine.

### Experiment on ResNet-152 with COVID-19

Due to the small amount of data, the network convergence speed is much faster. The number of training rounds is also much less than before. From the results, the accuracy rate reached 89.3%. For F1 score and AUROC value, they have reached good value.

### Comparison of Experimental Results

Table 1 compares the experimental results described in this article with the latest experimental results in the same field at home and abroad. However, not all the paper experiments use the same evaluation index, so if the paper does not mention the relevant items in the table, they will be replaced by blanks.

**Table 1.**
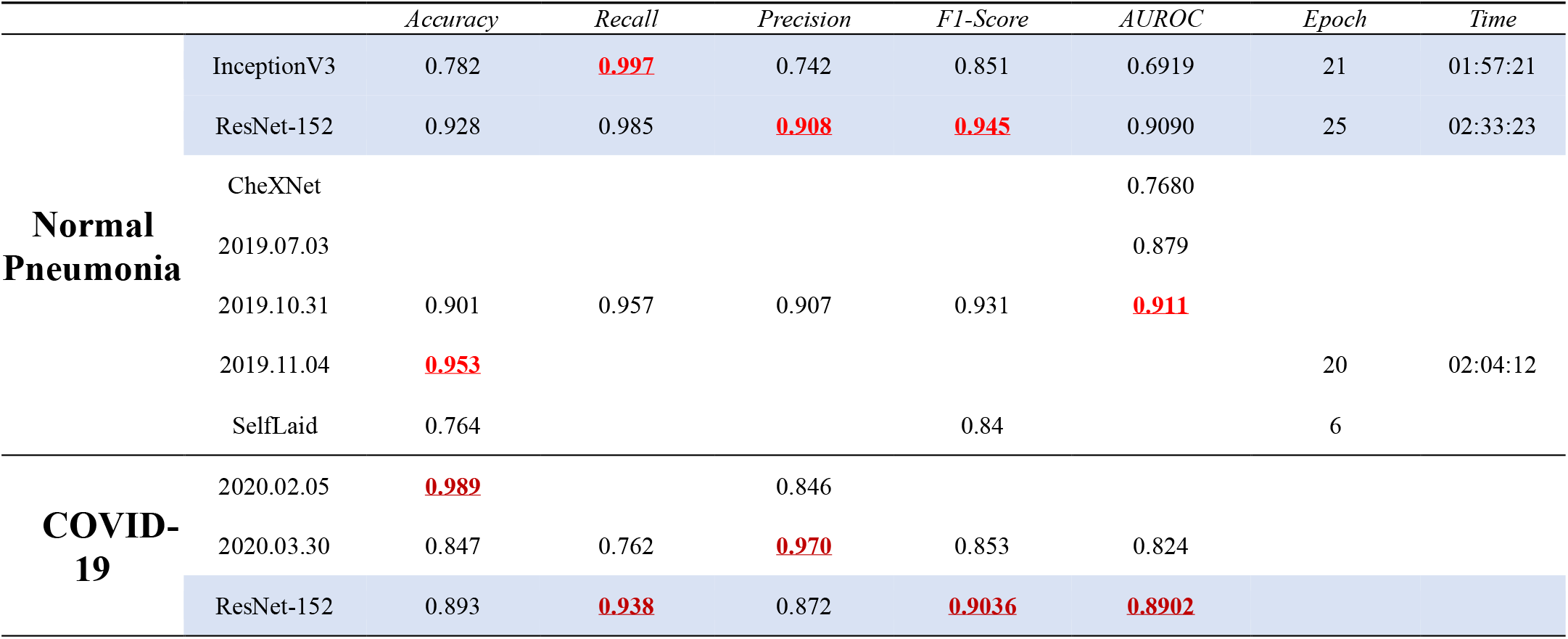
Results Comparison

The model represented by 2017.07.03 is derived from the *Using Deep Learning techniques for pulmonary thoracic segmentations and improvement of pneumonia diagnosis in pediatric chest radiographs* (Baisong et al., 2019). In practice, the image segmentation method was used first, and then the segmented images were classified. The focus of this paper is in of children with pneumonia diagnose. From the AUROC index perspective, this model has strong potential to replace human physicians (Baisong et al., 2019).

*Deep Convolutional Neural Network with Transfer Learning for Detecting Pneumonia on Chest X-Rays* was submitted on 2019.10.31 and was included in the book *Advances in Intelligent Systems and Computing* (Prateek et al., 2020). This paper combines deep learning with transfer learning and proposes a new model. From the experimental results of the paper, the performance of this model is excellent (Prateek et al., 2020). Transfer learning can apply the patterns learned in a specific field or task to different but related field problems, and complete the tasks in the target field with a limited amount of data. Therefore, in the future, in the face of some new diseases and limited data, artificial intelligence technology can still be used to diagnose the disease. In addition, it can be seen that there are still many existing methods in the field of artificial intelligence that can be used in medical imaging and further improve the accuracy of judgment.

The model represented by 2019.11.04 is derived from the paper *Pneumonia Radiograph Diagnosis Utilizing Deep Learning Network*, which was published in the 2019 IEEE 2nd International Conference on Electronic Information and Communication Technology (ICEICT) (Wesley et al., 2019). The experiments in this paper used the AlexNet model and Transfer Learning methods and obtained high accuracy in short-term training (Wesley et al., 2019; Krizhevsky, 2012).

As a model invented in 2012, nearly eight years have passed. During this time, the development of neural network models was very rapid, and models with higher practicality were also invented. Therefore, the application of neural network technology in medical imaging has excellent prospects.

The SelfLaid model is from the paper *Pneumonia Identification Using Chest X-Ray Images with Deep Learning*, which is a conference paper and is also included in the book *Advances in Intelligent Systems and Computing* (Nishit et al., 2019). The dataset used in this paper’s experiment is highly similar to the dataset used by us. According to the data disclosed in this paper, the accuracy of the model is not high enough (Nishit et al., 2019).

The experiment in *Deep learning-based model for detecting 2019 novel coronavirus pneumonia on high resolution computed tomography: a prospective study* did not use an X-ray dataset, but a CT dataset. This paper was submitted on MedRxiv and Arxiv on 2020.02.25, focusing on using deep learning techniques to determine COVID-19 (Jun et al., 2020). The author team of this paper is from mainland China. This paper shows the sensitivity of scholars in the field of artificial intelligence in China, and also reflects the new progress of Chinese artificial medical intelligence.

Some scholars from UCSD submitted a paper in Arxiv on 2020.03.30, which not only disclosed the dataset they collected, but also used a pre-trained DenseNet to train a convolutional neural network model and achieve an F1 of 0.85. COVID-19 is a worldwide problem, all scholars are working hard (Zhao, 2020).

Our neural network model, whether it is judging ordinary pneumonia or COVID-19, has shown excellent results. When the amount of data is sufficient, the trained neural network model can have better performances than the doctor. In the face of insufficient data, the trained neural network model can significantly improve the diagnosis efficiency while ensuring the accuracy of diagnosis. Our experimental results also prove this. Therefore, no matter whether it is a long-term disturbing disease like cancer or a sudden disease like COVID-19, we believe that artificial intelligence technology can play a key role in diagnosis and even further treatment.

## CONCLUSIONS

Genetic algorithms are similar to neural networks, and they all contain ideas that mimic the mechanisms of nature. When the internal mechanism of the target problem is not completely clear, a value close to the optimal solution can be obtained by the genetic algorithm. In the computer-related field, there are many cases of “non-zero is one”, so the application of the genetic algorithm is extensive and has strong robustness. In the experiments of this paper, by introducing the genetic algorithm, the dropout parameters adapted to the characteristics of the current network are obtained, thereby achieving better experimental results and improving the performance of the model.

Dropout, as an essential part of neural networks, has been proven effective in a large number of experiments, but how to make Dropout adapt to different models is a problem that needs to be further studied. The current Dropout algorithm stops the work of some neurons randomly. Could different neurons be given different weights, and random selection can be changed to weighted selection, which can maximize the effect of the Dropout function? The difficulty of this algorithm lies in the setting of weights. Our preliminary consideration is that the feature matrix of each passing neuron has specific characteristics mathematically. Is it possible to combine a matrix into a number after feature transformation, and give this number as a weight to the neuron? Our future work will focus on it.

As a typical bionic algorithm, genetic algorithm has excellent searchability, but its complexity is relatively high. Since there is one more step of preprocessing in the experiment than the usual method, it takes more time for finishing the automatic diagnostics. Therefore, our future research will make further efforts to optimize the bionic algorithms.

The completion of this paper coincided with the COVID-19 raging in the world (Yang, 2020). Through this epidemic, it could seem that there is still much space for improvement in China’s medical system. If artificial intelligence technology can assist doctors or even replace doctors in the diagnosis of medical imaging, it can significantly increase the number of patients per doctor per day. Perhaps when facing a similar crisis, our system can deal with it more calmly.

We expect artificial intelligence technology to be deeply involved in the medical system in the future, forming a complete medical process of “machine pre-diagnosis and provide basic information about the disease, professional doctors review and give treatment recommendations.” This can reduce not only the workload of doctors but also reduce the misdiagnosis rate of doctors so that improving the efficiency of the entire medical system. Also, in highly intelligent hospitals where “electronic doctors” are the first line, the number of doctor-patient contradictions may be significantly reduced, and the life safety of doctors may be protected to a large extent.

In conclusion, our experimental results prove that the Dropout module optimized by the genetic algorithm can play an important role in the neural network. Also, it can be seen that artificial intelligence technology has the potential to replace humans in the field of medical imaging completely.

## Data Availability

The source of the datasets are listed.

https://arxiv.org/abs/2003.13865

https://nihcc.app.box.com/v/ChestXray-NIHCC

## Notes

### Competing Interest Statement

The authors have declared no competing interest.

### Funding Statement

This work was supported by the National Natural Science Foundation of China (61075075,61110306140).

